# The epidemiology and clinical features of HIV and *Trypanosoma cruzi* (Chagas disease) co-infection: A systematic review and individual patient data analysis

**DOI:** 10.1101/2024.12.24.24319596

**Authors:** Natalie Elkheir, Jessica Carter, Catherine Dominic, Pat Lok, Temitope Fisayo, Melina Michelen, Barbara De Barros, Jaimie Wilson Goldsmith, Michael Butler, Amy Price, Anushka Mehotra, Laura Nabarro, Nadia Ahmed, Peter Chiodini, David A.J. Moore

**Affiliations:** Clinical Research Department, London School of Hygiene & Tropical Medicine, London, UK; Hospital for Tropical Diseases, University College London Hospitals, London, UK; UK Chagas Hub (https://www.uclh.nhs.uk/uk-chagas-hub); St George’s University of London, UK; Queen Mary’s University of London, UK; Norfolk and Norwich University Hospitals NHS Trust, UK; Anglia Ruskin University, Chelmsford, UK; University College London Hospitals NHS Trust, UK; School of Health Sciences, City University of London, London, UK; Royal Free London NHS Foundation Trust, UK; Imperial College Healthcare Trust, London, UK London; North West University Healthcare NHS Trust, London, UK; One Health Lewisham (GP Federation), London, UK; Central and North West London NHS Foundation Trust, London, UK

## Abstract

**Background:** Narrative descriptions of HIV and *Trypanosoma cruzi,* the causative agent of Chagas disease, co-infection exist in the literature but the breadth and depth of the data underlying these descriptions has not been previously thoroughly scrutinised and reactivation is poorly understood. The aim of this systematic review was to identify, synthesise and analyse the published literature on the epidemiology and clinical features of *T. cruzi* and HIV co-infection.

**Methods:** A systematic review of published literature on HIV and *T. cruzi* co-infection was conducted. Six international databases were searched: Medline, Embase, Global Health, Global Index Medicus, Web of Science and Scopus. Articles reporting on HIV and *Trypanosoma cruzi* co-infection, as defined by the authors, with no restrictions on study type, language or date of publication or reporting were included.

**Results:** 152 articles (62% case reports or series) were included which reported on 1,603 individuals with co-infection and 225 with presumed reactivation. Reported prevalence of co-infection varied greatly by region and setting of screening, from 0.1 to 1% in unselected populations, and was particularly high when screening inpatients known to have HIV for *T. cruzi* infection (26-48%). 83% of reactivations were reported in individuals with CD4<200 cells/mm^3^. CNS reactivation, typically presenting with meningoencephalitis and/or cerebral lesions, accounted for 68% of all published cases of reactivation. Myocarditis (accounting for 9% published reactivation cases) was less well characterised. Mortality of all reactivation cases was 59% (77% in those with CNS reactivation).

**Conclusion:** *T. cruzi* reactivation mainly affects those with untreated HIV and lower CD4 counts. CNS reactivation is the most common clinical picture and confers high mortality. Prompt recognition of reactivation and immediate initiation of trypanocidal therapy (with benznidazole or nifurtimox) is recommended. Increased education and better awareness of the risks of co-infection are needed, as is systematic screening of individuals at-risk.

## Introduction

Chagas disease is one of the most important zoonotic infectious diseases in Latin America and is increasingly being recognised as a public health problem in non-endemic settings within migrants from Latin America(1-3). It is caused by the protozoan parasite *Trypanosoma cruzi* and is endemic in the 21 countries of Central America, South America and Mexico. About 7 million people worldwide are estimated to be infected with *T. cruzi* and approximately 12,000 people die each year from clinical manifestations of Chagas disease(4). Infection is believed to be lifelong without treatment. The consequences of this chronic infection include progression to determinate cardiac or gastrointestinal end-organ disease, which affects approximately one third of individuals with *T. cruzi*. Chronic infection also poses an additional risk of parasite reactivation and replication in the context of immunocompromise such as immunosuppressant medication or HIV.

The first recognised description of *T. cruzi* reactivation in people living with HIV (PLHIV) was reported in Argentina in 1990, in a patient with central nervous system reactivation(5). Since then, many articles have reported high mortality rates associated with HIV/*T. cruzi* co-infection(6-8). A systematic review including articles published until 2010 identified 291 cases of co-infection published in the literature, and 100% mortality in untreated cases of reactivation(9). A narrative review published in 2021 described the epidemiology of co-infection, the clinical presentations most commonly associated with *T. cruzi* reactivation in PLHIV and highlighted the many gaps in knowledge(10). This systematic review will complement these earlier papers, through drawing together all international literature and publicly available data to provide an analysis of the current state of knowledge.

## Aims & Objectives

The aim was to systematically identify, synthesise and evaluate the evidence on the epidemiology and clinical features of HIV and *T. cruzi* co-infection, with a view to informing clinical and public health management, screening policy, and identify research priorities.

## Methods

A systematic review was conducted according to PRISMA guidelines(11), and registered with the International Prospective Register of Systematic Reviews (PROSPERO ID: CRD42020216125).

### Eligibility criteria

#### Inclusion criteria

This review includes articles reporting on HIV and *T. cruzi* co-infection, as defined by the authors, with no restrictions on study type, language or date of publication or reporting.

#### Exclusion criteria

Editorials and opinion papers were excluded, along with any articles not presenting primary data. Studies that included people with HIV and *T. cruzi* as a sub-group but did not present disaggregated data for any of the outcome measures of interest were excluded, unless corresponding authors were able to provide the required information (following two email attempts to contact authors). Articles where HIV and *T. cruzi* prevalence were reported separately (mainly with reference to blood bank screening and with no way to infer co-infection) were excluded. Systematic reviews and reviews were excluded, but citation tracking of any such publications was undertaken to identify additional publications not otherwise identified by the search strategy.

#### PICO question

Population: People of any age with *T. cruzi* infection/Chagas disease

Exposure: HIV co-infection, as defined by authors Comparison: N/A

Outcomes:

1. Prevalence of co-infection
2. Characteristics of patients and co-infection, including but not limited to:

a. Age
b. Sex
c. Country of birth/residence
d. CD4 count and HIV viral load
e. Clinical presentation
f. Setting of diagnosis and method of diagnosis (e.g. clinical, serological, parasitological)
g. Clinical investigations (including bloodwork, imaging and cerebrospinal fluid examination)
h. Treatment (antiretroviral, antiparasitic therapy and other)
i. Clinical outcomes: measures of morbidity and mortality

### Information sources

The following six databases were search on 1^st^ July 2022: Ovid Medline, Ovid Embase, Ovid Global Health, Global Index Medicus, Web of Science and Scopus. For any review articles identified, the listed references were screened, and citations of the articles were reviewed to identify any additional papers not captured by the original search strategy. Authors were contacted to request additional data where data was missing.

### Search strategy

Controlled subject headings and keywords of the following concepts were searched:

1) HIV

AND

2) Chagas disease or *Trypanosoma cruzi* infection There were no limits on language or date of publication.

### Selection process

Title and abstract screening of all records identified by the search strategy was carried out independently by two reviewers (NE and JC). Subsequent full-text screening of selected articles against the inclusion criteria was carried out independently by the same two reviewers, with all discrepant assessments resolved by consensus discussion. The Rayaan Systematic review data automation tool was used to organize articles and record screening decisions.

### Data collection process

Data were managed using Endnote for de-duplication of articles, Rayyan for screening, Microsoft Excel for data extraction and STATA version 18 for data analysis. The data extraction tool was developed in Microsoft Excel and piloted with input from the study team. Following this pilot assessment, the data extraction tool was split into two: one for articles that presented only aggregate-level data (e.g. prevalence studies) and one for articles with individual-level data (predominantly case reports and case series). Data extraction was performed independently by two reviewers (NE and either CD, PL, TF, MM, BdB, JWG, MB, AP or AM), who were fluent or native in the language of the article. Data extraction was checked by a third reviewer (JC) using the DeepL Pro (Cologne, Germany) online translation tool if necessary. Disagreements on interpretation of data to extract were resolved, and consensus reached, through discussion. Where important data was missing, the corresponding authors of the articles were contacted (twice via email) to request it.

### Data items

The following data (where available) was extracted from all articles: study design, aim, country of study, setting and context, inclusion and exclusion criteria, study population size (including prevalence of co-infection where reported) and characteristics, diagnostic criteria for HIV and *T. cruzi* infection, CD4 count, HIV viral load, clinical syndromes (signs and symptoms), investigations (including laboratory and radiological findings), treatments (antiretroviral, antiparasitic and others) and clinical outcomes (measures of morbidity and mortality).

### Study risk of bias assessment

The Joanne Briggs Institute Critical Appraisal Checklists (12) were used to assess for risk of bias in the studies included in the review. Two reviewers (NE and JC) assessed for risk of bias independently and reached consensus through discussion where disagreements arose. No formal measures to assess reporting bias or certainty in the body of evidence were taken, although these are qualitatively described.

### Effect measures

Prevalence of co-infection in the study population was extracted where available (for both HIV amongst subjects with *T. cruzi* infection and for *T. cruzi* infection amongst subjects with HIV), and the range of estimates were presented according to study setting. For studies reporting individual-level patient data, mortality was calculated as an overall proportion.

### Synthesis methods

Articles were analysed in two groups: those that presented only aggregate-level data, and those that presented any individual patient-level data. Articles that reported prevalence of co-infection were summarised in a table according to the setting (and context) of presentation. A meta-analysis of prevalence estimates was not performed due to the substantial heterogeneity among the different contexts.

For articles with individual-level data, due to heterogeneity in reporting of reactivation, the study team classified clinical presentations reported in articles into phenotypic groups (according to presence and/or likelihood of reactivation, described further below) by consensus. The individual patient analysis focussed on individuals with probable reactivation.

Continuous data for all patients were presented as the mean and standard deviation (if normally distributed) or median and interquartile range (if not), and comparisons between groups were performed using the Student’s t-test or Mann-Whitney *U* test respectively. The categorical data for all patients are presented as counts and frequencies, and comparisons were performed using the *χ*^2^ or Fisher’s exact test, as appropriate. All analyses were conducted using STATA version 18 and a *P* value of less than 0.05 was considered statistically significant.

## Results

### Study selection

Ovid Medline, Ovid Embase, Ovid Global Health, Global Index Medicus, Web of Science and Scopus were searched (as per methods section) on 1^st^ July 2022. Figure 1 outlines the process from article identification to inclusion.

**Figure 1.**
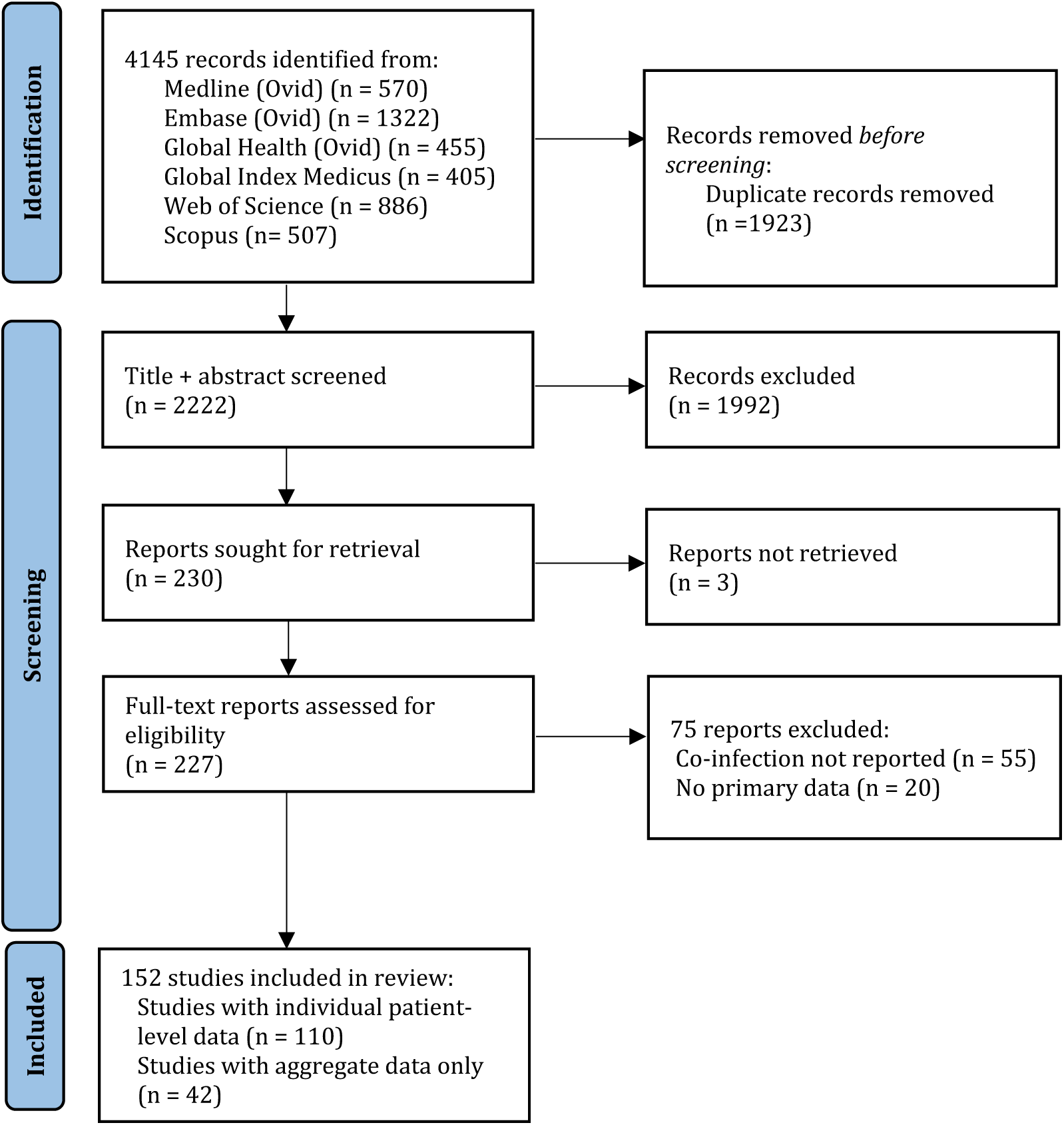
PRISMA (Preferred Reporting Items for Systematic reviews and Meta-Analyses) flow diagram outlining study selection. Database searches were performed up to 1^st^ July 2022.

### Study characteristics

152 articles were included in this systematic review, of which 131 (86%) were journal articles, 20 (13%) were conference abstracts or posters and one (1%) an online report. These articles consist of case reports (n=68, 45%), case series (n=26, 17%), cross-sectional studies (n=51, 34%), cohort studies (n=5, 3%) and case-control studies (n=2, 1%).

These 152 studies report on 1,603 individuals with HIV and *T. cruzi* co-infection (noting that, although we excluded duplicate patients reported across multiple articles in our individual level patient analysis where obvious, it is possible that some duplicates may have been missed where it was not clear from the articles).

121 studies were performed in endemic countries (Fig 2): Brazil (n=60, 39% of all included articles), Argentina (n=39, 26%), Colombia (n =7) Bolivia (n=6), Chile (n=5), Uruguay (n=1), Venezuela (n=2) and Mexico (n=1). Thirty studies were performed in non-endemic countries: Spain (n=14, 9%), US (n=13, 9%), Italy (n=3) and the UK (n=1).

**Figure 2.**
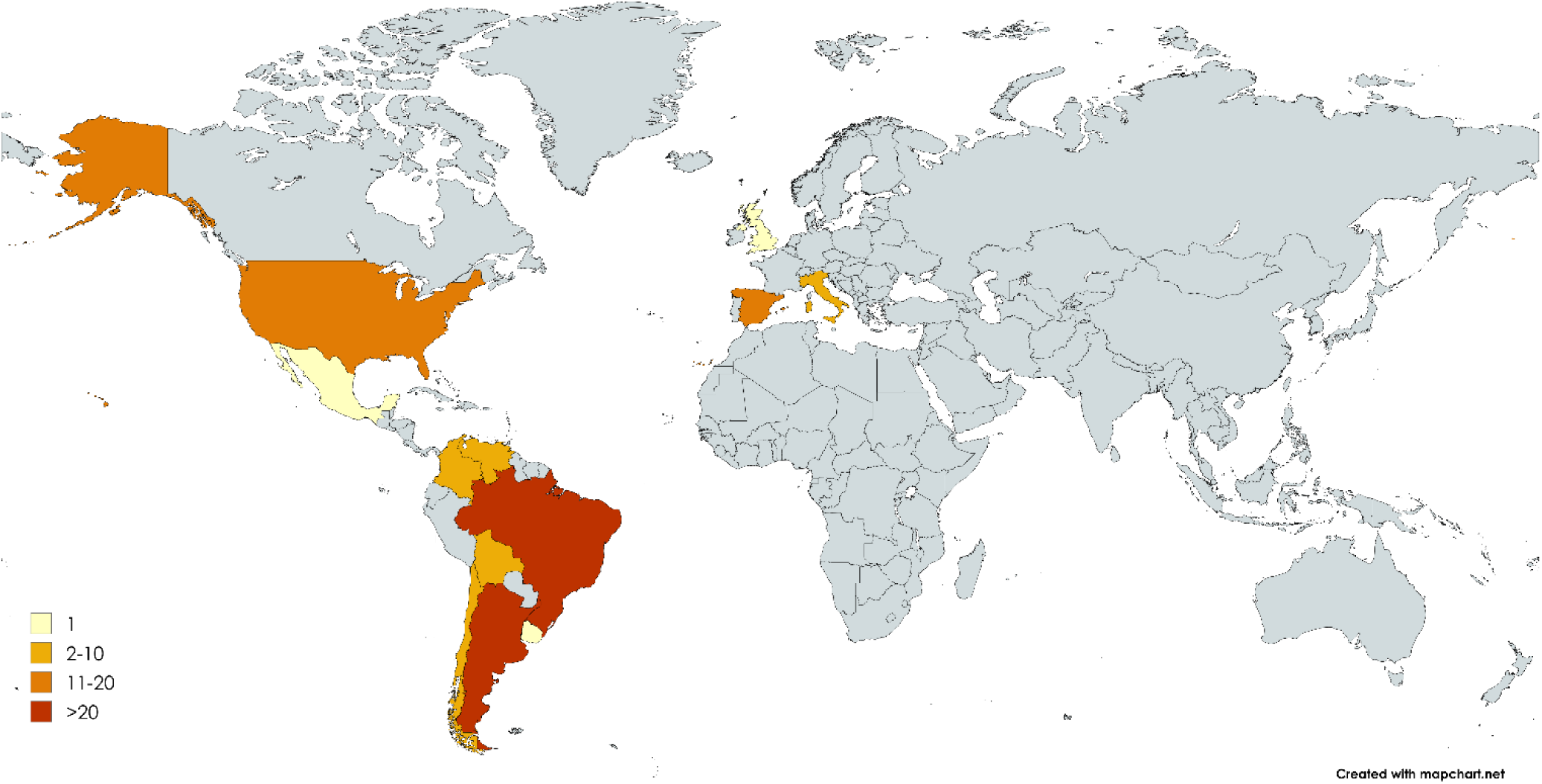
Number of articles published on HIV/T. cruzi co-infection by country setting

Table 1a outlines the characteristics of included articles that presented any prevalence data for HIV/*T. cruzi* co-infection, either *T. cruzi* infection amongst subjects with HIV or HIV infection amongst individuals with *T. cruzi* infection or both, or aggregate data only (i.e. no individual-level patient data).

**Table 1a.** Study characteristics and profile of HIV/*T. cruzi* co-infection in articles presenting prevalence of co-infection or only aggregate-data.

|  |  |  |  |  |  |
| --- | --- | --- | --- | --- | --- |
| **Author, year** | **Country** | **Study design** | **Population description** | **Co-infection/total P\*** | **Proportion** |
| **Screening people not known to have HIV or *T. cruzi* infection** |  |  |  |  |  |
| Carneiro-Proietti, 1998(22) | Brazil | Cross-sectional | Patients with haemophilia | 2/226 | 0.009 |
| Aguiar, 1999(23) | Brazil | Cross-sectional | First time blood donors to blood bank in Mato Gross do Sul State | 1/476 | 0.002 |
| Auger, 2002(13) | Argentina | Cross-sectional | Patients seen in emergency department in Buenos Aires (IPD** n=2) | 2/1680 | 0.001 |
| Bocanegra, 2014(14) | Spain | Cross sectional | Migrants from Chagas-endemic countries attending hospital in Barcelona | 4/416 | 0.01 |
| Birrer, 2021(24) | Brazil | Cross-sectional | Blood donors to clinic in Brazil | 1/296 | 0.003 |
| **Testing for *T. cruzi* infection in people with known HIV** |  |  |  |  |  |
| **Chagas-endemic countries** |  |  |  |  |  |
| Spina-França, 1988(25) | Brazil | Cross-sectional | PLHIV<sup>†</sup> with neurological presentations and CSF<sup>‡</sup> to hospital (IPD n=1) | 1/200 | 0.005 |
| Livramento, 1989(26) | Brazil | Cross-sectional | PLHIV with neurological presentations and CSF<sup>‡</sup> analysis (IPD n=2) | 2/170 | 0.012 |
| Torrealba, 1990(27) | Chile | Case-series | PLHIV with neurological presentation and brain biopsy (IPD n=1) | 1/4 | 0.25 |
| Reiche, 1996(28) | Brazil | Cross-sectional | PLHIV attending hospital in Londrina, Brazil | 4/181 | 0.022 |
| Lazo, 1998(29) | Brazil | Case-series | PLHIV with encephalitis (IPD n=15) | 15/22 | 0.682 |
| Oelemann, 2000(30) | Brazil | Cross-sectional | PLHIV with chronic diarrhoea (IPD n=4) | 4/95 | 0.042 |
| Scapellato, 2001(31) | Argentina | Cross-sectional | Seroprevalence of *T. cruzi* in PLHIV attending Buenos Aires Hospital | 8/184 | 0.043 |
| Rodrigues, 2005(32) | Brazil | Cross-sectional | PLHIV with and without *T. cruzi* infection, assessing cytokine serum levels | 18/28 | 0.643 |
| Scapellato, 2006(33) | Argentina | Cross-sectional | *T. cruzi* seroprevalence in PLHIV who inject drugs | 3/328 | 0.009 |
| Almeida, 2010(34) | Brazil | Cross-sectional | PLHIV at hospital (IPD n=20, 9 from cross-sectional study plus 11 from clinics) | 9/716 | 0.013 |
| Warley, 2010(35) | Argentina | Cross-sectional | PLHIV on ART<sup>§</sup> with malignancy or infection | 4/235 | 0.017 |
| Reimer-McAtee, 2014(36) | Bolivia | Cross-sectional | PLHIV presenting to hospital and use of PCR | 38/133 | 0.286 |
| Bowman, 2015(37) | Bolivia | Cross-sectional | PLHIV hospitalised in Santa Cruz | 37/157 | 0.236 |
| Benchetrit, 2016(38) | Argentina | Cross-sectional | *T. cruzi* seroprevalence in PLHIV + MSM , STI<sup>¶</sup> clinic attendees & IVDU<sup>#</sup> | 8/280 | 0.029 |
| Stauffert, 2017(39) | Brazil | Cross-sectional | Seroprevalence of *T. cruzi* in PLHIV in South West Brazil | 10/200 | 0.05 |
| Ignacio-Junior, 2018(40) | Brazil | Cross-sectional | PLHIV at Emilio Ribas Hospital | 2/240 | 0.008 |
| Dias, 2018(15) | Brazil | Cross-sectional | PLHIV seen in outpatient clinic in South West Brazil | 1/539 | 0.002 |
| Reimer-McAtee, 2021(16) | Bolivia | Cross-sectional | *T. cruzi* PCR in PLHIV in hospital in Cochabamba (IPD n=4, all with reactivation) | 32/116 | 0.276 |
| **Non-endemic countries** |  |  |  |  |  |
| Rodríguez-Guardado, 2009(21)<sup>††</sup> | Spain | Cross-sectional | *T. cruzi* screening in Latin American migrant PLHIV (IPD n=2) | 2/19 | 0.105 |
| Rodríguez-Guardado, 2010(41)<sup>††</sup> | Spain | Cross-sectional | *T. cruzi* screening in Latin American migrant PLHIV | 2/19 | 0.105 |
| Rodríguez-Guardado, 2012(42) | Spain | Cross-sectional | Migrant PLHIV attending hospital in Asturias | 2/57 | 0.035 |
| Llenas Garcia, 2012(43) | Spain | Cross-sectional | *T. cruzi* screening in Latin American migrant PLHIV in Madrid hospital (IPD n=3) | 4/154 | 0.026 |
| Salvador, 2013(44) | Spain | Cross-sectional | *T. cruzi* screening in migrant PLHIV | 5/126 | 0.04 |
| Ahmed, 2016(45) | UK | Cross-sectional | *T. cruzi* screening in Latin American PLHIV in sexual health clinic (IPD n=1) | 1/40 | 0.025 |
| Evans, 2018(20) | US | Cross-sectional | *T. cruzi* screening in Latin American migrant PLHIV (IPD n=2) | 2/179 | 0.011 |
| Rodari, 2022(46) | Italy | Cross-sectional | *T. cruzi* seroprevalence in Latin American migrant PLHIV | 5/389 | 0.013 |
| **Testing for HIV in people with known *T. cruzi* infection** |  |  |  |  |  |
| **Chagas-endemic countries** |  |  |  |  |  |
| Bisugo, 1998(47) | Brazil | Cross-sectional | Patients with known *T. cruzi* seen in hospital | 32/78 | 0.41 |
| Perez-Ramirez, 1999(18) | Brazil | Cross-sectional | Patients with known *T. cruzi* in four hospitals (IPD n=33) | 34/71 | 0.479 |
| Sartori, 2002(17) | Brazil | Cross-sectional | Patients with known *T. cruzi* | 29/110 | 0.264 |
| **Non-endemic countries** |  |  |  |  |  |
| Ramos, 2009(48) | Spain | Cross-sectional | Patients with known *T. cruzi* seen in three hospitals in Alicante | 1/67 | 0.014 |
| Escoin, 2011(49) | Spain | Cross-sectional | Patients with known *T. cruzi* seen at tertiary centre | 2/282 | 0.007 |
| Salvador, 2015(50) | Spain | Cross-sectional | Patients with *T. cruzi* in three hospitals in Catalonia (IPD n=12) | 12/1823 | 0.007 |
| Ramos-Rincon, 2021(51) | Spain | Cross-sectional | All patients discharged from a hospital with *T. cruzi* | 45/5022 | 0.009 |
| **Death certificates, registries & reports on co-infection** |  |  |  |  |  |
| Rocha, 1994(52) | Brazil | Cross-sectional | Autopsies of patients with HIV/*T. cruzi* co-infection | 23/23 | 1 |
| Martins Melo, 2012(8) | Brazil | Cross-sectional | Death certificates, all deaths in Brazil 1999-2007 | 74/8942217 | 0.0008 |
| Garcia-Pino, 2014(53) | Colombia | Cross-sectional | Autopsies and DNA analysis of PLHIV | 6/155 | 0.039 |
| Melo-Urbe, 2014(54) | Colombia | Cross-sectional | Autopsies of PLHIV and opportunistic infections | <20/155 | <0.13 |
| Gattoni, 2015(55) | Brazil | Cross-sectional | Autopsies of PLHIV with opportunistic infections (including Chagas reactivation) | 2/326 | 0.006 |
| Hernandez, 2017(56) | Colombia | Cross-sectional | Autopsies of PLHIV and opportunistic infections in Santander | 13/3497 | 0.0037 |
| Olivera, 2021(57) | Colombia | Cross-sectional | Death reports in Colombia of patients with known *T. cruzi* 1979-2018 | 95/3276 | 0.029 |
| Martins Melo, 2022(19) | Brazil | Cross-sectional | Death certificates all deaths in Brazil, 2000-2019 | 196/22663092 | 0.000009 |
| **Studies of known co-infection** |  |  |  |  |  |
| Braz, 2001(58) | Brazil | Cross-sectional | *T. cruzi* parasitaemia in immunocompromised patients | - | NA<sup>++</sup> |
| Sartori, 2007(59) | Brazil | Cohort | Observational cohort of known co-infection 1989-2005 | 53/53 | NA |
| Lages-Silva, 2009(60) | Brazil | Cross-sectional | *T. cruzi* genetic characterization in +/- HIV, +/- co-infection & +/- reactivation | 38/91 | NA |
| Scapellato, 2009(61) | Argentina | Cohort | Mother to child transmission of *T. cruzi* (3/94 mothers co-infected with HIV) | 3/94 |  |
| De-Freitas, 2011(62) | Brazil | Cross-sectional | PCR, blood culture and xenodiagnosis in patients with known co-infection | 32/91 | NA |
| Shikanai-Yasuda, 2014(63) | Brazil | Cohort | Parasitaemia in immunosuppressed patients, including co-infection | 43/43 | NA |
| Castro-Sesquen, 2016(64) | Bolivia | Case-control | Nanotechnology in urine in patients with HIV/*T. cruzi* co-infection | 20/39 | NA |
| Fernandez, 2015(65) | Argentina | Cross-sectional | Known HIV/*T. cruzi* co-infection with reactivation at Buenos Aires Hospital | 23/23 | NA |
| Tozetto-Mendoza, 2017(66) | Brazil | Cross-sectional | Cytokine producing T cells in Chagas, co-infection, HIV and healthy controls | 11/35 | NA |
| Fernandez, 2018**(67)** | Argentina | Case-series | Known HIV *T. cruzi* co-infection with neuro-reactivation in hospital | 7/7 | NA |
| Shikanai-Yasuda, 2021**(68)** | Multiple | Cross-sectional | Clinical profile and mortality of co-infection from multi-centre network | 241/241 | NA |

\* = Number with co-infection / total (at-risk) study population. \*\*Articles that reported any Individual patient data (IPD) are indicated, along with number of patients (n) with extractable data included in individual patient analysis and therefore table 1b.
† = People living with HIV, ‡ = Cerebrospinal fluid, § = Highly active antiretroviral therapy, ¶ = Men who have sex with men, ¶ = Sexually-transmitted infection, # = Intravenous drug user, †† = duplicated data therefore the article with more complete dataset included in IPD analysis, ‡‡ = not applicable

Table 1b outlines the characteristics of included articles that had any individual-level data which could be extracted (and therefore include in our individual patient data analysis).

**Table 1b.** Study characteristics of articles presenting individual patient date on HIV/*T. cruzi* co-infection.

| Author, year | Country | n | Author, year | Country | n | Author, year | Country | n |
| --- | --- | --- | --- | --- | --- | --- | --- | --- |
| **Endemic countries** |  |  | Lages-Silva, 2002(100) | Brazil | 1 | Simioli, 2016(135) | Argentina | 1 |
| Spina-França, 1988(25) | Argentina | 1 | Chirino, 2001(101) | Venezuela | 1 | Fica, 2017(136) | Chile | 1 |
| Livramento, 1989(26) | Brazil | 2 | Auger, 2002(13) | Argentina | 2 | Lopez, 2018(137) | Argentina | 1 |
| Del Castillo, 1990(5) | Argentina | 1 | Santos, 2002(102) | Brazil | 1 | Benchetrit, 2018(138) | Argentina | 9 |
| Torrealba, 1990(27) | Brazil | 1 | Rodrigues, 2002(103) | Brazil | 1 | Chalub, 2019(139) | Argentina | 1 |
| Ferreira, 1991(69) | Brazil | 1 | Sartori, 2002(104) | Brazil | 1 | Guidetto, 2019(140) | Argentina | 2 |
| Gallo, 1992(70) | Brazil | 1 | Antunes, 2002(105) | Brazil | 1 | Reimer-McAtee, 2021(16) | Bolivia | 4 |
| Oddo, 1992(71) | Chile | 2 | Marques De Brito, 2003(106) | Brazil | 2 | Fernández, 2021(141) | Argentina | 6 |
| Cardozo, 1992(72) | Uruguay | 1 | Madalosso, 2004(107) | Brazil | 1 | Muñoz-Calderón, 2022(142) | Argentina | 3 |
| Rosemberg, 1992(73) | Brazil | 1 | Da-Cruz, 2004(108) | Brazil | 3 | Marcon, 2022(143) | Brazil | 8 |
| Rocha, 1993(74) | Brazil | 1 | Auger, 2005(109) | Argentina | 8 | Bowman, 2022(144) | Bolivia | 4 |
| Libaak, 1993(75) | Argentina | 1 | Burgos, 2005(110) | Argentina | 1 | **Total no. patients in endemic settings\*** |  | **320** |
| Metze, 1993(76) | Brazil | 1 | Valergaa, 2005(111) | Argentina | 1 | **Non-endemic countries** |  |  |
| Nishioka, 1993(77) | Brazil | 1 | Corti, 2006(112) | Argentina | 1 | Gluckstein, 1992(145) | US | 1 |
| Solari, 1993(78) | Chile | 1 | Salcedo, 2006(113) | Mexico | 1 | Rivera J, 2004(146) | US | 1 |
| Freilij, 1995(79) | Argentina | 2 | Sartori, 2007*(59) | Brazil | 53 | Yoo, 2004(147) | US | 1 |
| Sartori, 1995(80) | Brazil | 1 | Burgos, 2008(114) | Argentina | 1 | Lury, 2005(148) | US | 1 |
| Freilij, 1995(81) | Argentina | 3 | Sica, 2008(115) | Argentina | 3 | Lambert, 2006(149) | US | 1 |
| Pimentel, 1996(82) | Brazil | 1 | Cordova, 2008(116) | Argentina | 15 | Rodríguez-Guardado, 2009(21) | Spain | 2 |
| Di Lorenzo, 1996(83) | Argentina | 2 | Diazgranados, 2009(117) | Colombia | 1 | Verdú, 2009(150) | Spain | 1 |
| Ferreira, 1997(84) | Brazil | 3 | De Almeida, 2009(118) | Brazil | 1 | Campo, 2010(151) | US | 1 |
| Montero, 1998(85) | Argentina | 2 | Moretta, 2009(119) | Argentina | 1 | Rodríguez-Guardado, 2011(152) | Spain | 1 |
| Iliovich, 1998(86) | Argentina | 1 | De Almeida, 2009(120) | Brazil | 2 | Llenas-Garcia, 2012(43) | Spain | 3 |
| Pacheco, 1998(87) | Brazil | 1 | Rodríguez, 2009(121) | Colombia | 1 | Angheben, 2013(153) | Italy | 1 |
| Cohen, 1998(88) | Argentina | 1 | Rocha, 2010(122) | Brazil | 1 | Mina, 2013(154) | Italy | 1 |
| Sartori, 1998(89) | Brazil | 18 | Lopez, 2010(123) | Chile | 1 | Yasukawa, 2014(155) | US | 1 |
| Sartori, 1998(90) | Brazil | 1 | Menghi, 2011(124) | Argentina | 1 | Salvador, 2015(50) | Spain | 12 |
| Lazo, 1998(29) | Brazil | 15 | Catay, 2010(125) | Argentina | 1 | Martinez-Ales, 2016(156) | Spain | 1 |
| Galhardo, 1999(91) | Brazil | 1 | Almeida, 2010(34) | Brazil | 20 | Alyemni, 2016(157) | US | 1 |
| dos Santos, 1999(92) | Brazil | 1 | Agosti, 2012(126) | Argentina | 2 | Ahmed, 2016(45) | UK | 1 |
| Filho, 1999(93) | Brazil | 1 | Bisio, 2013(127) | Argentina | 1 | Shah, 2017(158) | US | 1 |
| Pagano, 1999(94) | Argentina | 10 | Castrillon, 2013(128) | Argentina | 1 | Gomez, 2018(159) | US | 1 |
| Silva, 1999(95) | Brazil | 5 | Rotta, 2013(129) | Brazil | 1 | Evans, 2018(20) | US | 2 |
| Sartori, 1999(96)* | Brazil | 1 | Hernández, 2014(130) | Colombia | 1 | Multani, 2019(160) | US | 1 |
| Nisida, 1999(97) | Brazil | 4 | Spadafora, 2014(131) | Venezuela | 1 | Bosch-Nicolau, 2019(161) | Spain | 1 |
| Perez-Ramirez, 1999(18) | Brazil | 33 | Uliarte, 2015(132) | Argentina | 1 | Bustamante, 2020(162) | US | 1 |
| Concetti, 2000(98) | Argentina | 1 | Bucchieri, 2015(133) | Brazil | 1 | **Total no. patients non-endemic settings\*** |  | **38** |
| Corti, 2000(99) | Argentina | 1 | Castro-Sesquen, 2016(134) | Bolivia | 31 |  |  |  |
| Oelemann, 2000(30) | Brazil | 4 |  |  |  |  |  |  |

\*Cases duplicated in articles removed from total

### Risk of bias in studies

The majority of articles were case reports or case series (94 articles, 62%), which are particularly susceptible to selection bias and publication bias. Additionally, the analysis of individual patient data may have unknowingly counted patients twice where authors have published multiple series, further contributing to risk of bias.

All 152 papers were critically appraised using the design-appropriate Joanna Briggs Institute (JBI) critical appraisal tool. Seven articles (5%) were assessed as high quality (a score above 90%), 83 articles (55%) medium quality (score of 75-90%), and the remaining 62 articles (41%) as low quality (score <75%). The majority of the cases reports and case series were of low quality. The mean JBI score for all included articles was of low quality, at 67%.

### Results of individual studies

#### Proportion of study populations with co-infection

Table 1a outlines study characteristics of articles that presented prevalence of co-infection. In the five studies of testing of people not previously known to have either *T. cruzi* or HIV, the proportion found to have co-infection ranged from 0.1% in emergency department attendees in Argentina (Auger 2002)(13) to 1% in migrants from Chagas-endemic countries in Spain (Bocanegra 2014)(14).

Twenty-five studies reported testing for *T. cruzi* amongst people living with HIV (PLHIV). Excluding the nine studies where patients were explicitly selected based upon specific clinical presentations or risk groups (e.g. PLHIV with diarrhoea or neurological disease), the proportion testing positive for *T. cruzi* ranged from 0.2% in an outpatient setting in Brazil (Dias 2018)(15) to 28.6% in an inpatient setting in Bolivia (Reimer-McAtee 2021)(16).

Three studies, all from Brazil, reported testing for HIV in patients with known *T. cruzi* infection in hospital settings, amongst whom co-infection prevalence ranged from 26.4% (Sartori 2002)(17) to 47.9% (Perez-Ramirez 1999)(18).

Martins Melo et al. reported that both Chagas disease and HIV were mentioned on 196 death certificates in Brazil (out of 22,663,092) between 2000 and 2019(19).

Thirteen studies reported on testing of migrants from Chagas-endemic countries living in non-endemic settings In the eight studies reporting testing for *T. cruzi* infection in PLHIV the proportion ranged from 1.1% in primary care in the US (Evans 2018)(20) to 10.5% in Spain (Rodriguez-Guardado 2009)(21).

#### Clinical features of co-infection

One cross-sectional retrospective multicentre study of co-infection (Shikanai-Yasuda, 2021)(68) reported on 241 patients with co-infection (87% from Brazil), of whom 60 were cases of reactivation. In these 60 patients with reactivation, 35 (58%) presented with meningoencephalitis, 10 (17%) with myocarditis and eight (13%) with both. 31 (52%) had CD4 counts below 200 cells/mm^3^ at presentation. Mortality overall was 67%, higher in those with meningoencephalitis (77%), who on average had lower CD4 counts compared to myocarditis (in whom mortality was 50%). This study did not report individual-level patient data so is not included in our analysis below, however most of the individuals with reactivation referred to in this article have been published in separate case series so are captured in our individual patient analysis.

110 studies reported individual-level patient data on clinical features or outcomes for a total of 357 patients with HIV and *T. cruzi* co-infection (once obvious duplicate patients reported across multiple articles were excluded). Study characteristics are summarised in table 1b. Demographics of patients with HIV/*T. cruzi* co-infection are summarised in table 2.

**Table 2.** Characteristics of 357 patients reported with co-infection (from 110 studies reporting individual-level patient data)

|  |  |  |
| --- | --- | --- |
| **Age, median (IQR)** |  | 36 (30-44) |
| **Sex** |  |  |
|  | Female | 85 (35%) |
|  | Male | 158 (65%) |
|  | Not reported | 114 |
| **Country of presentation** |  |  |
|  | Brazil | 180 (50%) |
|  | Argentina | 88 (25%) |
|  | Bolivia | 39 (11%) |
|  | Spain | 21 (6%) |
|  | US | 14 (4%) |
|  | Chile | 5 (1%) |
|  | Other (with <5 patients per country) | 10 (3%) |
|  | Not reported | 0 |
| **Country of birth** |  |  |
|  | Brazil | 143 (52%) |
|  | Bolivia | 55 (20%) |
|  | Argentina | 49 (18%) |
|  | Chile | 5 (2%) |
|  | El Salvador | 5 (2%) |
|  | Honduras | 5 (2%) |
|  | Other (with <5 patients per country of birth) | 14 (5%) |
|  | Not reported | 81 |
| *Denominator for given percentages is total data available (excluding missing data).* |  |  |

#### Definition of co-infection and reactivation

All 357 patients had laboratory-confirmed *T. cruzi* and HIV co-infection. There was a lack of an internationally standardised definition of *T. cruzi* reactivation in PLHIV and significant heterogeneity in the reporting of co-infection, with different clinical and laboratory parameters adopted historically and geographically. Therefore, for the purposes of this systematic review, individual cases were assigned one of three (broad) clinical phenotypes (Fig 3).

**Figure 3.**
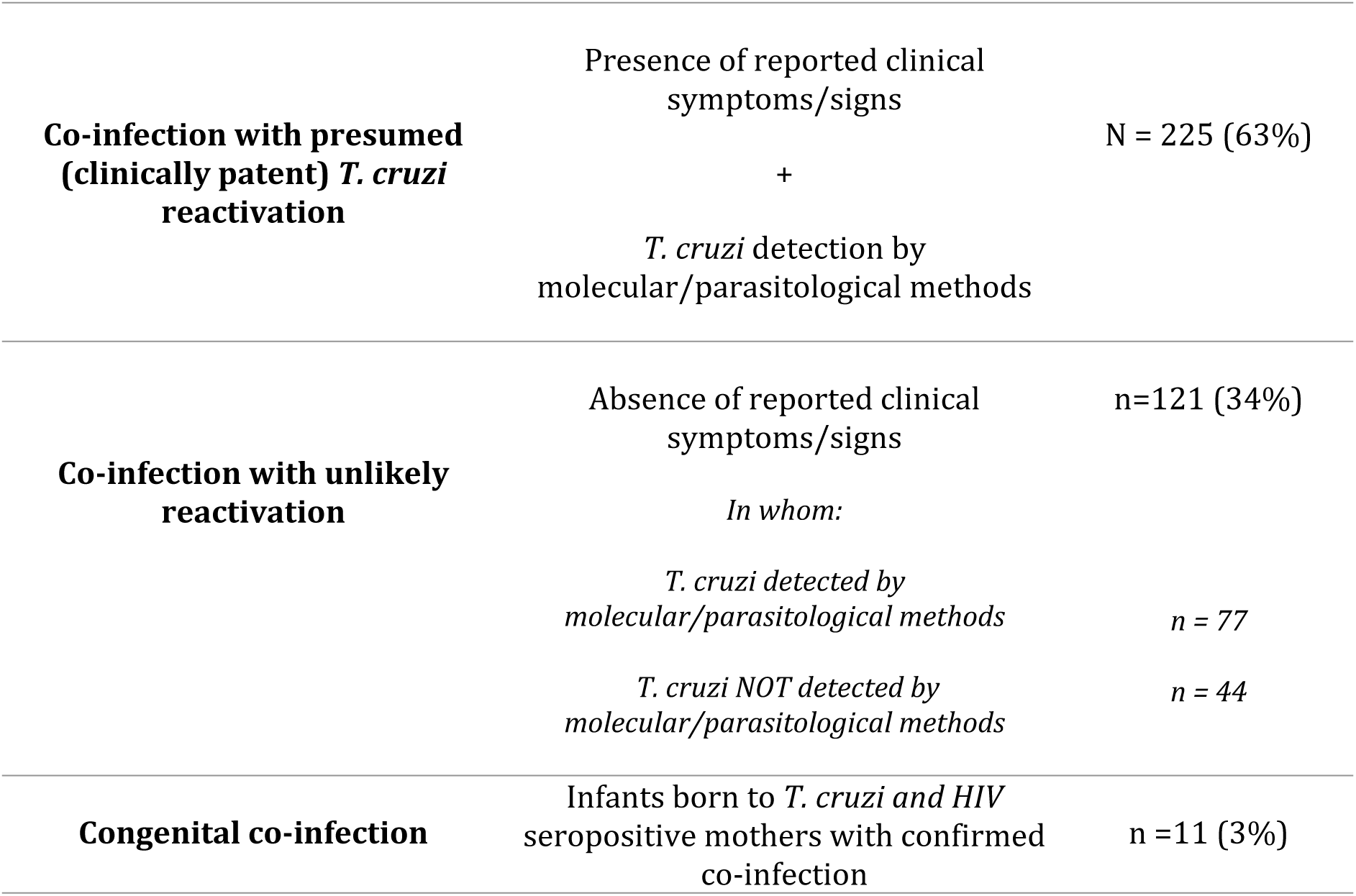
Clinical characterisation of 357 patients with HIV and *T. cruzi* co-infection

#### Individual patient analysis of *T. cruzi* reactivation

From here onwards, only the data from the 225 individuals who met our aforementioned ‘presumed *T. cruzi* reactivation’ criteria (Fig 3) are reported. The 225 subjects determined as having ‘probable reactivation’ had a median age of 36 (IQR 30-43). 116 (52%) were male, 49 (22%) female, and sex was not reported in 60 (27%) cases. 210 (93%) cases presented in endemic settings (50% of whom in Brazil), and 15 (7%) in non-endemic settings.

#### HIV and Trypanosoma cruzi status

156 (69%) cases were known to have HIV prior to presentation, HIV was unknown at the point of presentation in 52 (23%) patients and was not recorded in 17 (8%) cases. 37 (16%) cases of reactivation occurred in patients on antiretroviral therapy (ART) (none of whom were HIV virally suppressed in the seven cases where viral load was reported), 81 (36%) in ART-naïve cases and this data was missing in 107 (48%) cases. HIV viral load was reported in 30 cases of reactivation (median 179,976, IQR 44,000 – 409,000, range 4,700 – 1,800,000 copies/mL), none of whom were virally suppressed. 68 (30%) cases of reactivation were known to have *T. cruzi* infection prior to presentation, this was unknown in 102 (45%) patients and not recorded in 55 (24%) cases. No cases of reactivation were noted to have previously received trypanocidal treatment with benznidazole or nifurtimox, but one case had received ketoconazole (published in 1999)(91).

#### Clinical presentation of reactivation

Table 3 summarises the HIV and *T. cruzi* investigation findings of all cases of reactivation and Table 4 the presenting features.

**Table 3.** HIV and *T. cruzi* investigation findings of 225 patients with presumed reactivation.

| **Investigations, units**<br>**(n result available/total)** |  | **Number of patients** |
| --- | --- | --- |
| **CD4 count, cells/mm<sup>3</sup> (115/225)** |  |  |
|  | Mean (SD) | 114 (134) |
|  | Median (IQR) | 63 (25-177) |
|  | 0-99 | 73 (63%)* |
|  | 100-199 | 23 (20%)* |
|  | 200-499 | 19 (17%)* |
|  | >=500 | 3 (3%)* |
| **HIV viral load, copies/mL (30/225)** |  |  |
|  | Mean (SD) | 306,359 (408,608) |
|  | Median (IQR) | 179,976 (44,000-409,000) |
| ***T. cruzi* parasite detectable by:** |  | **No. patients (%)** |
|  | Blood microscopy | 76 (33%)<sup>†</sup> |
|  | Blood culture | 42 (19%)<sup>†</sup> |
|  | Xenodiagnosis<sup>‡</sup> | 63 (28%)<sup>†</sup> |
|  | Blood PCR | 28(12%)<sup>†</sup> |
|  | CSF microscopy | 68 (30%)<sup>†</sup> |
|  | CSF PCR | 9 (4%)<sup>†</sup> |
|  | CSF culture | 1 (<1%)<sup>†</sup> |
|  | CSF serology | 14 (6%)<sup>†</sup> |
|  | Biopsy | 55 (24%)<sup>†</sup> |
|  | Brain | 40 (17%)<sup>†</sup> |
|  | Skin | 4 (2%)<sup>†</sup> |
|  | Other | 11 (5%)<sup>†</sup> |

\*Denominator for given percentages is total data available for this variable (excluding missing data). <sup>†</sup>Denominator for given percentages is total number of patients (225). <sup>‡</sup>63 positive xenodiagnoses come from 15 papers, all in Brazil or Chile. PCR = polymerase chain reaction. CSF = cerebrospinal fluid

**Table 4.** Clinical presentation of 225 patients with presumed reactivation.

|  |  |
| --- | --- |
| Presentation | Number of patients (%) |
| **Fever** | 64 (28%) |
| **Neurological** |  |
| Focal deficit | 74 (33%) |
| Weakness | 65 (29%) |
| Headache | 56 (25%) |
| Seizures | 41 (18%) |
| Cognitive deficit | 25 (11%) |
| Loss of consciousness | 19 (8%) |
| Neck stiffness | 17 (8%) |
| Speech disturbance | 11 (5%) |
| Visual disturbance | 9 (4%) |
| Tremor | 2 (1%) |
| Any neurological presentation | 153 (68%) |
| **Gastroenterological** |  |
| Weight loss | 34 (15%) |
| Diarrhoea | 15 (7%) |
| Vomiting | 13 (6%) |
| Dysphagia | 8 (4%) |
| Any gastroenterological presentation | 37 (16%) |
| **Cardiorespiratory** |  |
| Shortness of breath | 12 (5%) |
| Cough | 8 (4%) |
| Palpitations | 1 (0.4%) |
| Chest pain | 1 (0.4%) |
| Any cardiorespiratory presentation | 20 (9%) |
| **Skin lesions** | 6 (3%) |
| *Denominator for percentages is total number of patients (225)* |  |

40 (18%) individuals with reactivation were reported as acutely unwell (clinical shock – as defined by authors or systolic blood pressure <90 mmHg, altered consciousness or requiring critical care).

### Central nervous system reactivation

Meningoencephalitis and/or cerebral space occupying lesions accounted for 68% of the cases of reactivation, with presentation with focal neurological deficit, headache or seizures most commonly reported, in 33%, 25% and 18% respectively.

Median CD4 counts in reactivation with CNS involvement were lower than those without CNS involvement (46 versus 123 cells/mm^3^, Wilcoxon rank-sum z statistic 4.52, p<0.0001).

95% of CNS reactivation cases had a CD4 count under 200 cells/mm^3^; 77% were under 100 cells/mm^3^.

#### Cerebrospinal fluid examination

93 (41%) patients with reactivation had a lumbar puncture (LP), of whom 85 had neurological symptoms and/or abnormal neuroimaging findings reported suggestive of CNS reactivation. The LP was reported as normal in four cases (none of whom had symptoms or neuroimaging suggestive of CNS reactivation) and abnormal in 89 cases (85 of whom had neurological symptoms and/or abnormal neuroimaging findings reported suggestive of CNS reactivation and four of whom did not). *T. cruzi* was detectable in the cerebrospinal fluid in 79 cases (93% of presumed CNS reactivation cases, table 3). Figure 4 displays the distribution of cerebrospinal fluid (CSF) white cell counts, protein and glucose, for patients with CNS reactivation.

**Figure 4.**
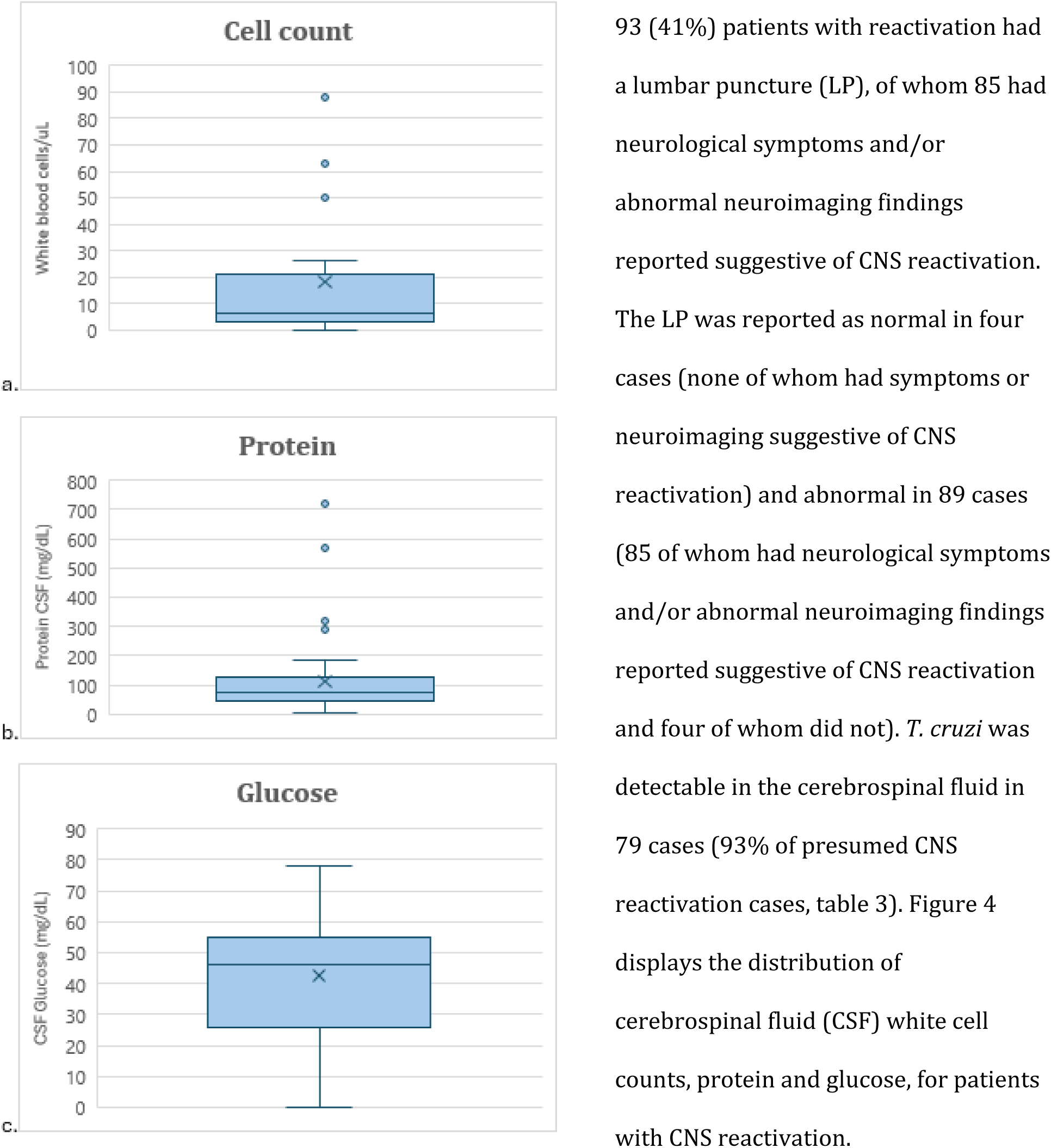
Box and whisker plots displaying the cerebrospinal fluid examination findings (a. white cell count, b. protein and c. glucose) in PLHIV and *T. cruzi* CNS reactivation.

Of 44 reported CSF cell counts, the cell count was greater than five white blood cells/µL in 50% of cases (greater than 10 in 46%, greater than 20 in 25%, greater than 50 in 14% cases) and, when reported, was predominantly lymphocytic.

According to a normal protein CSF reference range of 15–60 mg/dL, of 43 reported protein values, 25 (58%) were raised (45% greater than 80, 31% greater than 100, 10% greater than 200).

According to a normal glucose CSF reference range of 50–80 mg/dL, of 37 reported values, 22 (59%) reported low glucose and 15 (41%) reported normal glucose.

#### Neuroimaging

123 patient shad neuroimaging: 104 patients (46%) had a CT brain, 48 (21%) an MRI and 29 (13%) had both. In six symptomatic patients. In six symptomatic patients, neuroimaging was reported as normal (all six patients had an LP, five of which were reported – all abnormal, including three which had *T. cruzi* detectable by microscopy in the CSF). 54 (52% of those with neuroimaging) patients had a single lesion, and 60 (58%) patients had multiple (2 or more) lesions. 46 (44%) lesions were reported as ring-enhancing, eight (8%) reported as located in white matter, three (3%) in grey matter (and location not stated in the majority of reports). Cerebral oedema was present in 54 (52%), with a mass effect in 37 (36%).

### Cardiac reactivation

Myocarditis was reported in 20 cases (9% of all reactivation reports) and was a common finding on autopsy (13 out of 34 cases, 38%). The median CD4 count of the 11 cases for whom CD4 count was reported was higher (median 136, [IQR 102-264]) than for those with CNS reactivation (Wilcoxon rank-sum z = -2.4, p<0.05).

#### Cardiorespiratory investigations

In the absence of prior data on cardiac function, attribution of reported cardiac abnormalities to *T. cruzi* reactivation cannot be substantiated, nevertheless the reported findings (for all patients with presumed reactivation) were as follows: ECG was reported as abnormal in 51 (22%) cases (supraventricular tachycardia = 4, ventricular tachycardia = 1, right bundle branch block = 21, left bundle branch block = 9, atrioventricular block = 11, bradycardia = 1, ventricular ectopics = 6, ST changes = 1, prolonged QT = 1). Chest x-ray was reported as abnormal in 19 (8%) cases (cardiomegaly = 9, pneumonia = 5, infiltrates = 5, effusion = 4). Echocardiography was reported as abnormal in 24 (11%) cases (systolic dysfunction = 16, thrombus = 1, valve disease = 4, pericardial effusion = 5, dilatation = 7).

### Other clinical presentations

Other clinical presentations of *T. cruzi* reactivation in PLHIV were less frequently reported, and included erythema nodosum in three cases(59, 96, 137), and pleuritis(124), peritonitis(86), cervicitis (98) and panniculitis(137) in one case each.

### Opportunistic infections

Concurrent additional opportunistic infections were commonly reported (n=86, 38%). Cryptococcosis was reported in six cases (3%), herpes simplex virus in eight cases (4%), tuberculosis in 13 cases (6%), cytomegalovirus in 14 cases (6%), pneumocystis pneumonia in 19 cases (8%), candidiasis in 25 cases (11%) and toxoplasmosis in 28 cases (17%). In the cases of concurrent cryptococcosis, cytomegalovirus and toxoplasmosis, CNS involvement was usually inferred but rarely well described.

### Management

Anti-retroviral treatment (ART) was poorly reported. ART was initiated in 65 (29%) patients at some point (65/225, 131 had missing data). Delaying initiation of ART was explicitly noted in 16 (7%) cases. 137 patients (61%) were treated for reactivation disease with trypanocidal drugs (120 with benznidazole and 17 with nifurtimox). Adverse effects from trypanocidal drugs were reported in 25 patients (18% of those treated), with 12 of those 25 patients having to stop trypanocidal treatment because of this. The mean trypanocidal treatment duration was 58 days (standard deviation 94, range 1 – 730). 56 (25%) patients were treated for toxoplasmosis.

Steroid therapy was given in 34 (15%) cases. Data on ART choice and timing, as well as secondary (trypanocidal) chemoprophylaxis were lacking. Immune reconstitution inflammatory syndrome (IRIS) was reported (as probable, rather than confirmed) in two cases (136, 157), both of whom died.

### Outcomes

Overall mortality was 59% (132 patients died out of 225 with reactivation). Nine (4%) survived with morbidity (of which seven had neurological sequelae, one had congestive cardiac failure and one not described). Forty-nine patients (22%) fully recovered whilst outcome data was missing for 35 (16%) patients.

Overall mortality in patients with CNS reactivation (77%) was higher than those without (49%) CNS reactivation (x^2^ 14.45, p<0.001) Amongst those with data available on both ART at presentation and clinical outcome, there were 12 deaths in 36 people reported to be on ART at presentation (33% mortality), compared to 55 deaths in 77 people known to not be taking ART at presentation (71% mortality).

Time to death was reported in 78 cases, amongst whom it occurred at a median of 30 days after presentation (IQR 9 - 84 days, range 0-4,200). Median time to death in cases of CNS reactivation was (statistically significantly) shorter (21 days, IQR 6-57) than for those without CNS reactivation (92 days, IQR 41-600, Wilcoxon rank sum z = 3.3, p<0.01).

In univariate analyses, age, sex and CD4 count were not statistically significant predictors of death, but neurological presentation was (x^2^ 14.45, p <0.001).

### Autopsies

38 autopsies were reported in people known to have HIV/*T. cruzi* co-infection (34 in cases of presumed reactivation). The macro- and microscopic patterns of necrosis, haemorrhage and inflammation of cerebral lesions, and direct observation of *T. cruzi* parasites, were described in 15 articles(29, 59, 69, 73, 74, 77, 91, 92, 99, 107, 111, 130, 131, 157, 158). Carditis (mostly myocarditis with amastigotes in muscle cells with infiltrates) was reported in 12 articles(71, 73, 74, 77, 80, 84, 91, 96, 97, 118, 130, 146). Nests of *T. cruzi* amastigotes with inflammation, necrosis and haemorrhage in the adrenal glands were reported in one article(103). Myositis of the extra-ocular muscles was reported in one article(92).

### Maternal and congenital co-infection

Eight articles reported maternal and/or congenital co-infection(50, 61, 79, 81, 97, 126, 127, 129). Scapellato(61) assessed *T. cruzi* vertical transmission rates in 94 *T. cruzi* seropositive mothers (three of whom were co-infected with HIV). All three neonates born to co-infected mothers had congenitally acquired *T. cruzi* (versus 10 out of 91, 10.9%, of those born to HIV negative mothers). Similarly Nisida(97) reported *T. cruzi* vertical transmission rates in 58 *T. cruzi* seropositive mothers: congenital *T. cruzi* transmission occurred in both infants born to the two HIV co-infected mothers compared to two out of 56 (4%) born to HIV negative mothers.

The outcomes of eight cases of co-infection in pregnant women (of which three were reactivation, all survived) were reported(43, 59, 97, 127). This included four cases published by Sartori et. al in which three resulted in congenital transmission. All three neonates were symptomatic with CNS disease (and two out of three neonates died) (59). One case report of *T. cruzi* CNS reactivation in a pregnant woman with co-infection reported the use of benznidazole at 32 weeks in successfully treating the woman; congenital infection did not occur (127).

Eleven congenital cases of co-infection were reported in six articles (50, 79, 81, 97, 126, 129), of whom nine were symptomatic, with meningoencephalitis (n=5), anaemia (n=3) and/or hepatosplenomegaly (n=3), and six died (of which three had neurological involvement and two with sepsis).

## Discussion

This systematic review provides the most comprehensive collation and overview of published evidence on the epidemiology and clinical manifestations of HIV and *Trypanosoma cruzi* co-infection and reactivation. Although our review includes a large body of evidence (152 articles published over 34 years, reporting on 1,603 patients in 12 countries), the majority of published articles were of poor quality with missing data on key variables. As outlined by Clark et al. in a 2021 narrative review on Chagas disease in people with HIV(10), we found significant heterogeneity in the way *T. cruzi* reactivation is reported across articles. The lack of any internationally recognised and standardised definition of reactivation hindered the interpretation of our results. To aid future research and guideline development, we recommend clearer reporting of clinical reactivation (in which trypanocidal treatment benefit is proven) versus parasitological/molecular reactivation in the absence of clinical symptoms (in which more research is needed to understand and guide monitoring and treatment regimens).

44 studies were identified that reported prevalence of *T. cruzi* and HIV co-infection (or proportion co-infected in selected populations), ranging from less than 0.0001% recorded co-infection on death certificates in Brazil between 2000 and 2019(19) to 48% when testing for HIV in inpatients with known *T. cruzi* infection in Brazil in 1999(18). A particularly high-risk group for co-infection was inpatients in endemic areas, especially individuals presenting with neurological symptoms (25, 26, 28). Despite advances in access to antiretroviral therapy(163, 164) reported in the literature and downward trending incidence of *T. cruzi* infection, the prevalence of co-infection reported in endemic hospital settings remained high in the more recent publications, for example 24% and 38% of hospitalised patients with known HIV screened for *T. cruzi* in two different hospitals in Bolivia in 2015 and 2021 respectively(16, 37). In one study from Argentina published in 2016, other HIV risk groups with high *T. cruzi* seroprevalence were men who have sex with men, STI clinic attendees and intravenous drug users, with an overall co-infection prevalence of 3%(38). Our findings support the recommendation for universal co-infection screening for those known to have either HIV or *T. cruzi* in endemic settings, ideally at the first point of contact with healthcare providers.

Co-infection prevalence in studies performed in non-endemic settings varied more widely, from 1-2% when screening Latin American migrants with HIV for *T. cruzi* infection in Italy, the US and the UK (20, 45, 46), to 3-11% in Spain (21, 43, 44). Some study authors attributed lower than expected prevalence to poor uptake of testing, especially amongst groups born in higher-endemicity countries. Low awareness of Chagas disease amongst at-risk migrants, as well as healthcare professionals in non-endemic countries, has been reported in the literature as a potential barrier to screening uptake (165-168). We recommend educational interventions, which have successfully been implemented, to improve uptake of screening in these settings(169), whilst acknowledging that the demographic profile of migrant communities from Latin America at highest risk of *T. cruzi* infection may not overlap substantially with that of migrant populations at highest risk of HIV infection.

This review identified very little evidence on maternal and congenital HIV-*T. cruzi* co-infection, and this is recommended as a priority area for future research. The eight published articles on this topic suggest high rates of congenital *T. cruzi* infection amongst neonates born to mothers with co-infection (compared to mothers with *T. cruzi* alone). The articles also suggest very high neonatal mortality associated with co-infection.

Risk of *T. cruzi* reactivation in immunosuppression was assessed in another recent systematic review, which reported a pooled cumulative incidence of reactivation of 17% (95% CI 8-29%) in PLHIV(170), which was lower than other contexts of acquired immunosuppression such as transplant recipients. In our review, we felt the level of heterogeneity between both study contexts and populations, as well as the different interpretations of what constitutes reactivation, plus the limited follow-up time in any prospective studies (which were lacking altogether) impeded a potential meta-analysis of risk of reactivation. More research is needed to understand the true risk of reactivation (including the effect of trypanocidal drugs), with a clearly defined study population and definition of reactivation, prospective outcome measurement, and sufficient sample size and follow-up duration.

This systematic review’s individual-patient analysis focused on 225 cases with probable reactivation. The majority of these patients were born in and presented to healthcare systems in Brazil, with a small representation of patients additionally from Argentina and Bolivia, and a small number of migrants from Bolivia, El Salvador or Honduras presenting in the US or Spain. Two thirds of the cases of presumed reactivation were reported in articles published since the year 2000 (22% since 2010), demonstrating that *T. cruzi* reactivation in the context of HIV is not a phenomenon confined to the pre-ART era and is still relevant to modern clinical practice.

Reactivation was mostly reported in patients with a CD4 count below 200 cells/mm^3^ (83% of patients), 63% were lower than 100. In our review, we found that CNS reactivation accounts for two thirds of the total cases of reactivation and is associated with lower CD4 counts (only 5% cases occurred in those with CD4 counts over 500 cells/mm^3^). Our findings on the clinical features of *T. cruzi* CNS reactivation are similar to those of a recently published review on this topic which found most (but not all) cases of CNS reactivation to occur in individuals with CD4 counts below 200 cells/mm^3^, a similar proportion of patients presenting with seizures and motor deficits, and a similar mortality overall (74%)(171). Our systematic review is more comprehensive, in that a more exhaustive search strategy was utilised, thus identifying more cases of reactivation. HIV viral load was underreported (only 30 cases reported this), nonetheless all 30 cases showed detectable viral loads. As has been previously reported(10), this review found CNS lesions from *T. cruzi* reactivation to be mainly (but not exclusively) ring-enhancing, and cerebral oedema was commonly reported. Expert groups with significant clinical experience in this area have reported that *T. cruzi* lesions tend to occur in white or subcortical matter, by contrast with Toxoplasma which may preferentially affect the thalamus and basal ganglia (172), however this was not clearly substantiated from our systematic review. As there are reports (n=6) of normal neuroimaging in *T. cruzi* CNS reactivation, CSF examination (ideally including *T. cruzi* PCR) is recommended in cases of sufficient clinical suspicion, even if imaging is reported as unremarkable.

Neuroimaging can be normal in *T. cruzi* meningoencephalitis, and where available the use of PCR in CSF to enhance sensitivity is recommended. Alongside the other main differential diagnoses for PLHIV presenting with neurological features (toxoplasmosis, primary CNS lymphoma, progressive multifocal leukoencephalopathy, tuberculosis and cryptococcosis for example), we recommend that *T. cruzi* should be considered in anyone who has spent time in any of the 21 Chagas-endemic countries of South America, Central America or Mexico.

We found a smaller proportion of cardiac reactivation (namely myocarditis) – fewer than 10% of published cases of reactivation, than expert review articles have suggested (30-40%)(172). As has previously been postulated, the burden of Chagasic cardiac reactivation is difficult to quantify as it may go unrecognised if not severe, that is, it could be attributed to chronic determinate disease. Additionally non-fatal cases are less likely to be published.

Robust data on management and outcomes of *T. cruzi* reactivation was lacking, although the lack of published reports of immune reconstitution inflammatory syndrome (IRIS) suggest this is not a common feature. However, it is possible that IRIS is not well described in these cases due to patients dying before IRIS manifested. Expert guidelines suggest waiting until at least three weeks of treatment with benznidazole (or nifurtimox) has been received prior to starting ART(172). Although the evidence-base is lacking, expert guidelines also recommend secondary chemoprophylaxis for patients who have experienced reactivation due to *T. cruzi*/HIV co-infection when CD4 level is below 200 cells/mm^3^ (but do not recommend primary prophylaxis)(172).

The main limitation of the body of evidence presented in this systematic review stems from the biases (namely selection bias and publication bias) to which case series are susceptible. Severe and fatal cases are more likely to be published, and the proportion of reactivation presenting with neurological symptoms may be overrepresented as they are more likely to be attributed to *T. cruzi* reactivation. One retrospective multicentre study of co-infection was identified, which reported on 241 patients with co-infection, mainly from Brazil, and it is a further limitation of our review that we could not extract individual level patient data from this study to include in our analysis (however it is thought most of the cases of reactivation were published elsewhere so have been captured in our search).

## Conclusion

Most evidence on HIV/*T. cruzi* co-infection and reactivation comes from case series, which are susceptible to selection and publication biases and therefore mainly of low quality. Evidence on maternal and congenital co-infection is particularly lacking. From the published literature, *T. cruzi* reactivation mainly affects those with untreated HIV and lower CD4 counts. CNS reactivation is the most common clinical picture, typically causing meningoencephalitis and cerebral lesions, and confers high mortality. Prompt recognition of reactivation and immediate initiation of trypanocidal therapy (with benznidazole or nifurtimox) is recommended. Screening for co-infection should be implemented in endemic settings (for *T. cruzi* in those with HIV and vice versa) and for migrants from at-risk areas in non-endemic settings. Educational initiatives are needed to increase healthcare professionals’ awareness of Chagas disease, in particular of the risk of reactivation in HIV and other forms of immunosuppression.

## Registration

This systematic review was registered on Prospero (ID: CRD42020216125, available from: https://www.crd.york.ac.uk/prospero/display_record.php?ID=CRD42020216125)

## Data availability statement

All relevant data are within the manuscript or available in a data repository.

## Author summary

The consequences of infection with both HIV and *Trypanosoma* cruzi (the parasite which causes Chagas disease) has been written about in the literature since the 1990s. In this review article, the evidence on co-infection is systematically identified, summarized and critiqued. Most evidence comes from small case reports or series of individual patients and is not of high quality. The main risk of co-infection is reactivation in the central nervous system, when the *T. cruzi* parasite replicates, causing meningoencephalitis. This usually occurs when HIV is not well-controlled and is associated with a high risk of death. Prompt recognition of co-infection, and antiparasitic treatment of *T. cruzi* can mitigate this risk.

